# The Lasting Effects of the Pandemic: A Time Series Analysis of First-time Pediatric Speech Delays

**DOI:** 10.1101/2023.05.18.23290122

**Authors:** Brianna M. Goodwin Cartwright, Peter D Smits, Sarah Stewart, Patricia J Rodriguez, Samuel Gratzl, Charlotte Baker, Nicholas Stucky

**Affiliations:** Truveta, Inc. Bellevue, Washington

## Abstract

Given the profound effects of the COVID-19 pandemic on the way individuals interact, we sought to understand if there was an increase in pediatric first-time speech and language delay diagnoses. We identified children under five years of age with a first-time speech delay diagnosis between January 1, 2018 and February 28, 2023, in Truveta Data. We calculated the monthly rate of first-time speech delay diagnoses per children with an encounter within the last year and no previous speech delay diagnosis. The Seasonal-Trend decomposition using LOESS (STL) method was used to adjust for seasonality. We also compared the difference in means between the 2018/2019 and 2021/2022 time periods. Significant increases in the mean of rates between 2018/2019 and 2021/2022 exist for the overall population and each age strata (p<0.001). Likely the causes of these trends are multifaceted and future research is needed to understand the specific drivers at play.

---

Nearly 8% of children have a communication or speech and language disorder.[1] During the COVID-19 pandemic lifestyle changes affected the way children live and communicate, including (but not limited to) stay-at-home measures, increased mask wearing, and decreases in both pre-school attendance and in-person pre-school options.[2] One Uruguayan study found motor and cognitive losses for children during the pandemic compared to a control population.[3]

Little data exists from the US about the effects of the pandemic on children, specifically understanding speech delays. We are interested to see if there is a change in first-time speech and language delay diagnoses before and after the pandemic.

## Methods

We identified children under five years of age with encounters between January 1, 2018 and February 28, 2023, in Truveta Data. Truveta Data consists of electronic health records provided by member US healthcare systems (representing ambulatory centers, hospitals, imaging centers, and clinics and medical centers), that were deidentified by expert determination in accordance with the HIPAA Privacy Rule.[4–6]

We identified encounters associated with first-time speech delay diagnosis defined using diagnostic codes (**Appendix**). We calculated the monthly rate of first-time speech delay diagnoses per child with an encounter within the last year and no previous speech delay diagnosis. We looked at the under-five population overall and stratified by age (0-1, 1-2, 2-3, 3-4, and 4-5 years of age).

We used seasonal-trend decomposition using LOESS (STL) to create a seasonally adjusted time series of well-child visits. We also compared the difference in means between the 2018/2019 and 2021/2022 time periods; 2020 was not included due to variation in care throughout the year.

## Results

We identified 2,463,511 patients under 5-years old who had an encounter between January 2018 and February 2023 and no previous speech delay diagnosis; of these, 3.6% had a first-time diagnosis within this period (**Table 1**). Throughout the study period the two-year-old patients had the highest rate of first-time diagnoses, followed by the one-year-old patients (**Figure 1**).

**TABLE 1.**
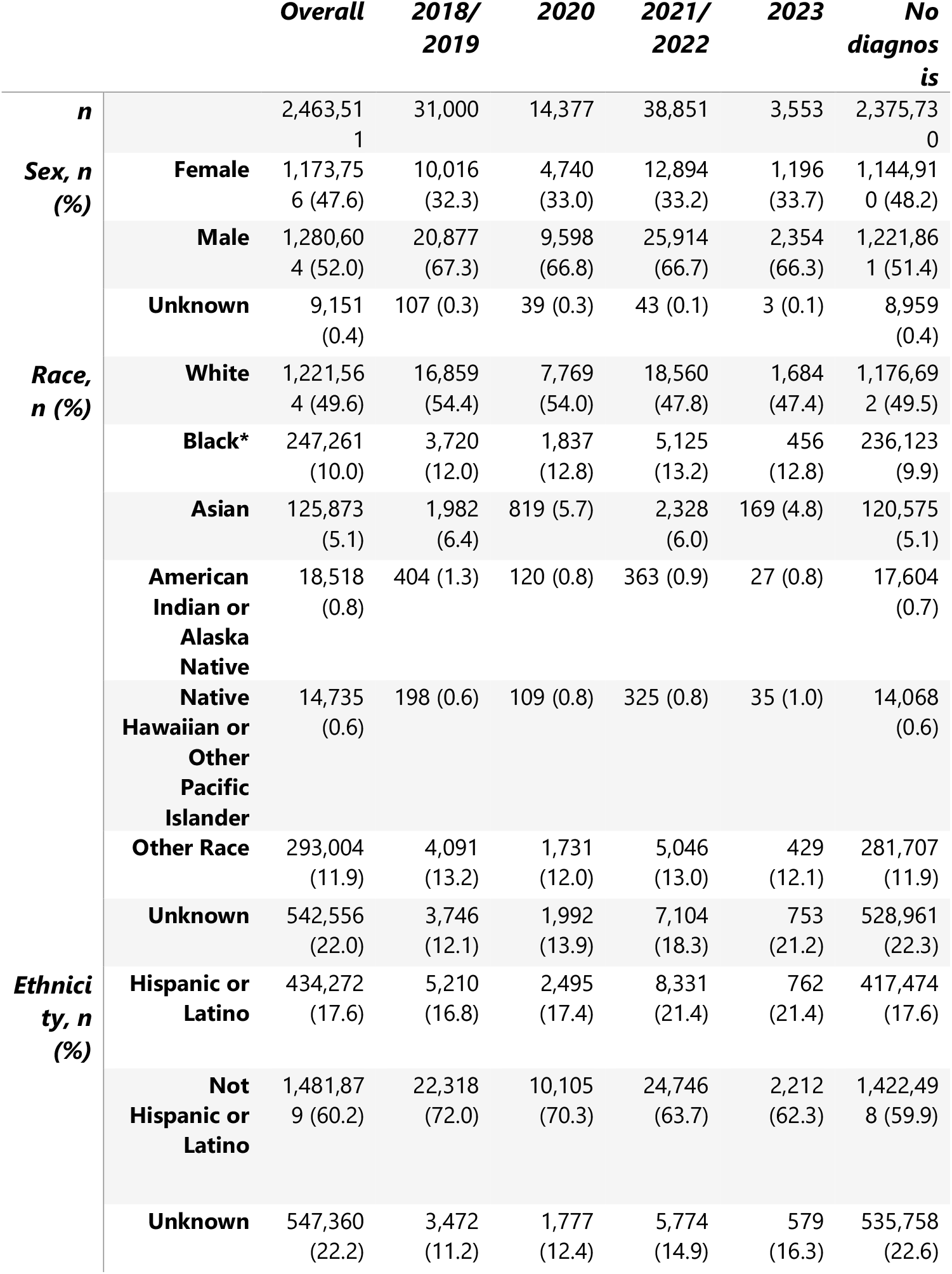
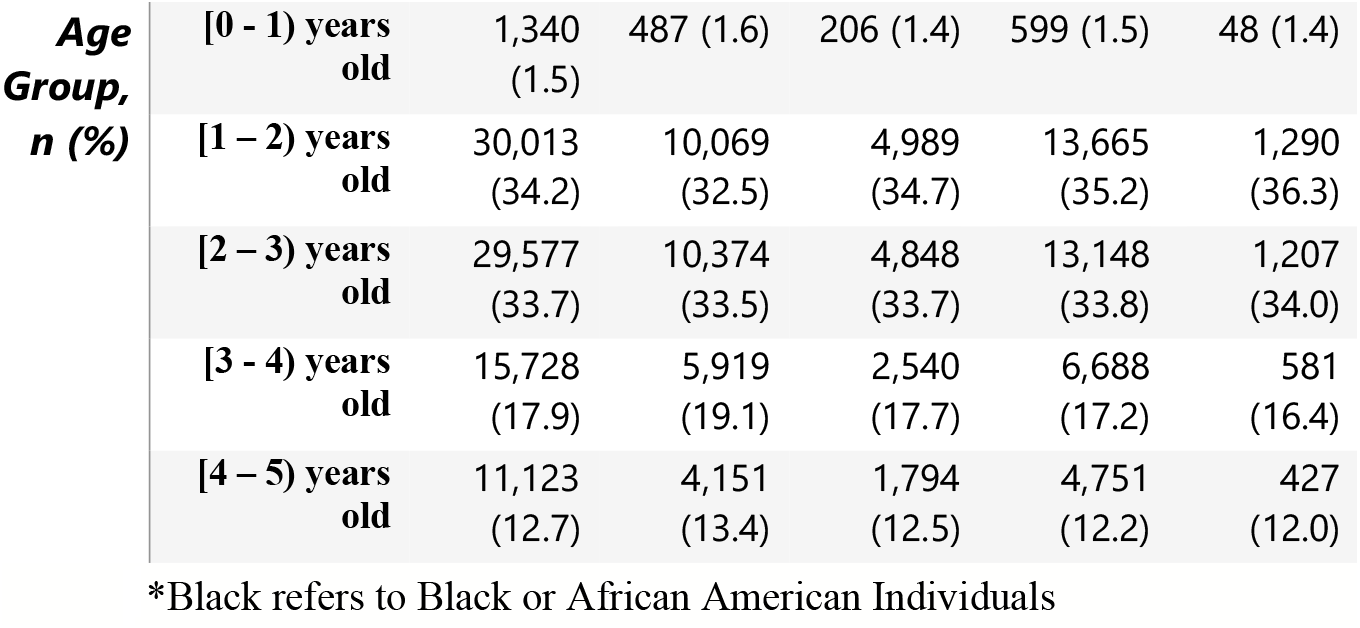
Demographics of population based on the year of speech delay diagnosis.

**FIGURE 1.**
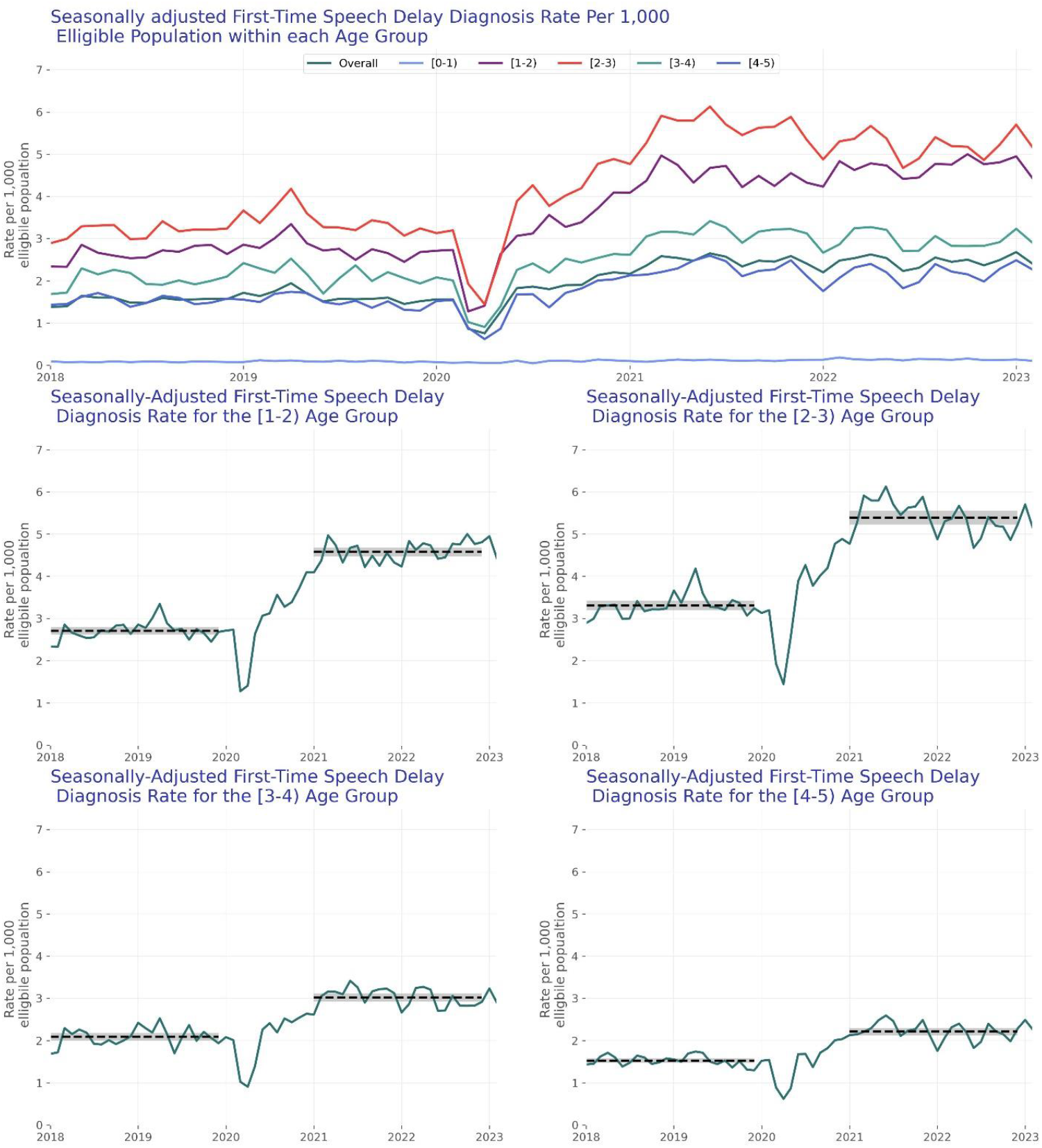
The seasonally adjusted first-time diagnosis rate of speech delays for each age group. The top panel shows overall and all age groups. The 4-lower graphs show each age group with rates above 1. The green line represents the trend, the dashed black line represents the 2- year mean, and the shaded grey regions are the 95% confidence intervals.

Significant increases in the estimated rates between 2018/2019 and 2021/2022 exist for the overall population and each age strata (p<0.001). Overall, we see a significant increase of 1.55x the rate of first-time speech diagnosis between 2018/2019 and 2021/2022 (2018/2019 rate: 1.58 [95% confidence interval: 1.53, 1.63], 2021/2022 rate: 2.45 [2.40, 2.51], p<0.001). We saw the largest increase in the patients who were one-year-old, where the rate increased by 1.69 times (2018/2019 rate: 2.71[2.62, 2.80], 2021/2022 rate: 4.58[4.47, 4.69], p<0.001). The patients who were two-years-old also increased by 1.63 times (2018/2019 rate: 3.31[3.19, 3.43], 2021/2022 rate: 5.39[5.23, 5.56], p<0.001).

## Discussion

We found a significant increase in the rate of first-time speech delay diagnosis for all age groups studied. The most pronounced increases occurred for the one- and two-year-old age groups, potentially due to regular screenings at these ages. Likely the causes of these trends are multifaceted and future research is needed to understand the specific drivers at play. Factors may include fewer social interactions, psychological distress, or a reduction in speech propagation while wearing masks.[7]

This study did not analyze severity of speech delay, therapy initiation or effectiveness; however, potentially amplifying the effects of these results, decreases in therapy effectiveness during the pandemic have been seen.[8] We also did not study a causal link between speech delays and any variables (such as mask wearing). Only diagnoses within Truveta health systems were available; school or private practice-based diagnoses were not included. Age during months without an encounter was estimated using the age at previous encounters. Despite these limitations, to our knowledge, these data are the first to show an increase in speech delay diagnoses in the years following the pandemic.

## Data Availability

The data used in this study is available to all Truveta subscribers and may be accessed at studio.truveta.com.

https://www.truveta.com/

## Appendix Diagnostic codes used to define speech delays.

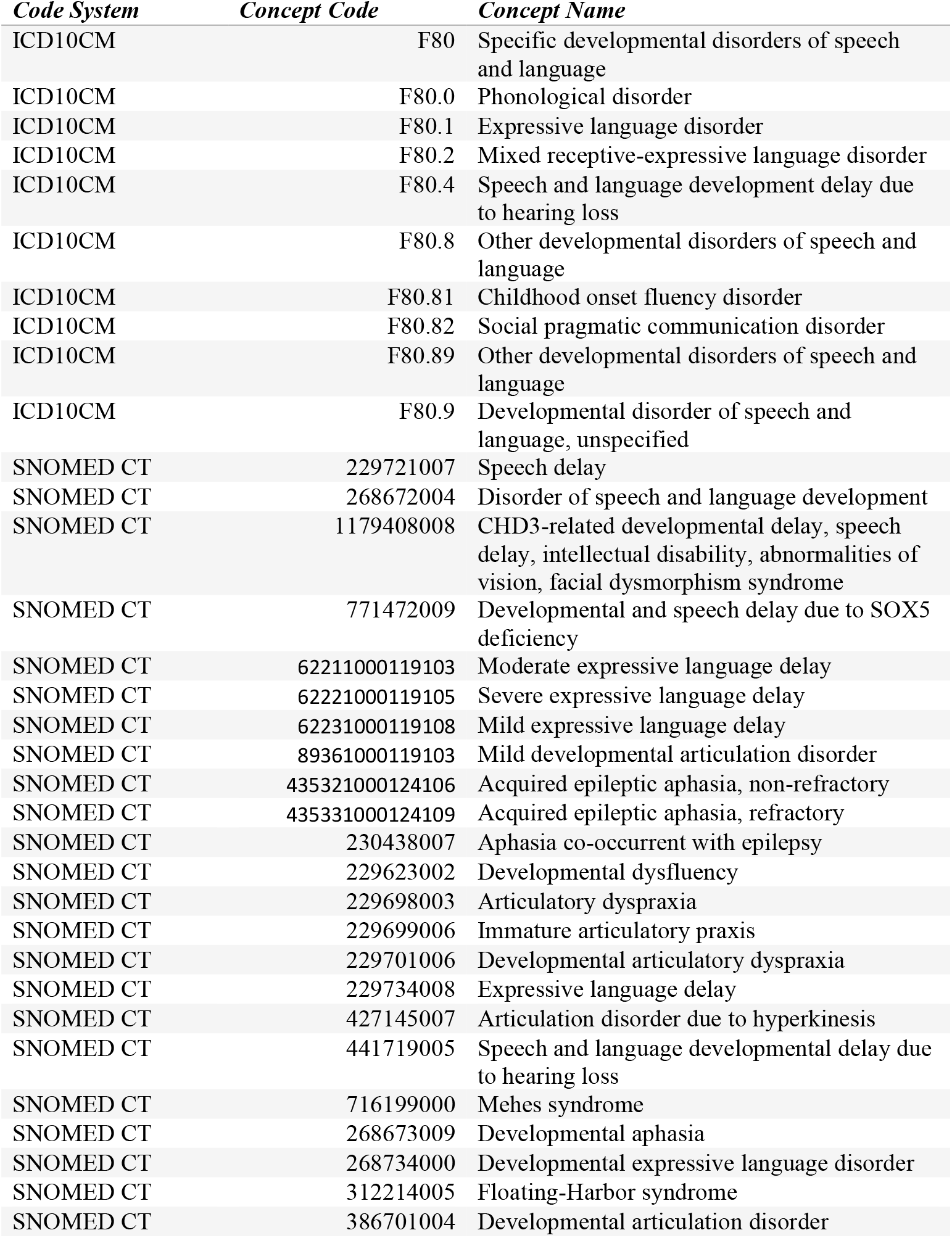
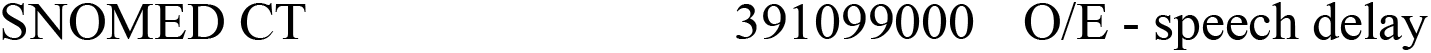

## Institutional review board statement

This study performs analysis of de-identified electronic health records (EHR) data accessed via Truveta Studio. Truveta Studio only contains data that has been de-identified by expert determination in accordance with HIPAA Privacy Rule, and therefore this study was exempt from Institutional Review Board approval.

